# Provision of medical Same Day Emergency Care services within the UK: analysis from the Society for Acute Medicine Benchmarking Audit

**DOI:** 10.1101/2024.10.13.24315407

**Authors:** C Atkin, M Perrett, M Holland, T Cooksley, T Knight, R Varia, C Subbe, DS Lasserson, E Sapey

## Abstract

**Aim:** To evaluate current provision of medical Same Day Emergency Care (SDEC) services within the UK, and current utilisation of these pathways in the assessment of unplanned medical attendances.

**Design:** Survey data was used from the Society for Acute Medicine Benchmarking Audit (SAMBA), including anonymised patient-level data collected annually using a day of care survey

**Setting:** Hospitals accepting unplanned medical attendances within the UK, 2019-2023. Participants: 34,948 unplanned and 4,342 planned attendances, across 188 hospital sites

**Results:** 29.8% of unplanned medical attendances received their initial medical assessment within SDEC services, with the proportion increasing over time. 82.4% of patients assessed in SDEC services were discharged without overnight admission. Assessment in SDEC services was less likely in male patients, patients with frailty, and older adults (all p<0.005).

Selected operational standards for SDEC delivery were met in 64-91% of hospitals. Most hospitals (82%) accepted referrals from emergency department triage and 63% accepted referral directly from the paramedic team. 38% of hospitals did not use a recognised selection criteria to identify suitable patients for SDEC and only 8% used a criteria designed to identify patients suitable for discharge. Overall, 34.7% of medical attendances discharged without overnight admission received their medical assessment in locations other than SDEC.

**Conclusions:** Medical SDEC provides assessment for one third of patients seen through acute medicine services. Although the proportion of patients assessed within SDEC is increasing, further innovation and improvements are needed to ensure appropriate patients access this service.

**Strengths and limitations of this study:**

-Medical same day emergency care (SDEC) has been widely adopted in the UK to deliver care to patients without overnight hospital admission, however there is limited evidence guiding development of this service.
-This study compares hospital-level data describing SDEC service structure and processes, and patient level data for over 35,000 patient attendances at 188 hospitals in the UK.
-This is the largest evaluation of medical SDEC to date and demonstrates an increase in the use of SDEC for medical patients nationally.
-The participation rate was higher amongst hospitals in England compared to the other three UK nations, which may limit generalisability.

## Introduction

Acute hospital services continue to experience increasing pressure, with more than 500,000 unplanned emergency admissions each month in England alone.(1) The majority of emergency admissions are due to medical conditions, which are assessed and managed through multiple pathways within acute services. The standard model of care within the UK is for patients requiring admission to an inpatient medical bed to be initially assessed and managed within an acute medical unit (AMU). However many patients can be assessed, treated and discharged without an overnight stay, through medical same day emergency care (SDEC) services.(2,3)

SDEC services and their UK counterparts, previously known as ambulatory emergency care (AEC), aim to provide timely assessment, investigation and treatment of emergency conditions without the need for overnight admission.(2,4) By reducing pressure on inpatient services and minimising the risks associated with hospital admission, SDEC should benefit individual patients and the healthcare service as a whole.(5,6) As a result, the increased utilisation of SDEC in all acute hospitals is a key ambition within the NHS Long Term Plan, which suggests that one in three patients should be discharged same-day, with improvements in this metric driven by increased utilisation of SDEC services.(7) This increase may be facilitated by appropriate identification of patients suitable for treatment within SDEC services, and by condition-specific ambulatory management pathways.(8)

Despite the widespread adoption of SDEC as a model of care within acute services, there remains considerable variation in the provision of SDEC services between hospitals, with limited evidence evaluating the impact of SDEC provision on patient flow, pathways through acute services or patient care itself.

We aimed to describe the current structure of medical SDEC services within the UK, using data from the Society for Acute Medicine Benchmarking Audit (SAMBA), a day-of-care survey that evaluates the structure and performance of acute medicine services within the UK. We also aimed to assess variation in the utilisation of the SDEC pathway for planned and unplanned medical attendances, evaluating patient factors associated with management through same day services.

## Methods

Data were collected through SAMBA. Participation in SAMBA is voluntary and open to all hospitals accepting unplanned admissions to acute and/or general internal medicine; community hospitals were excluded. Multiple hospital sites can register from each Trust/Health Board or equivalent, using the Society for Acute Medicine (SAM) website. Local approvals were obtained by individual sites, including Caldicott Guardian approval. Health Research Authority approval has been granted to allow secondary analysis on non-identifiable data (REC 21/HRA/4196). The full protocol for SAMBA is available online.(9) Study data were collected and managed using the CaseCapture database (NetSolving) for 2019-2021 and REDCap electronic data capture tools hosted at University of Birmingham for 2022-2023.(10)

Each round of SAMBA includes patient-level data for all medical admissions over a single 24-hour period on the penultimate Thursday of June (with the exception of winter SAMBA in January 2020), in conjunction with a unit-level organisational survey describing acute service structure. Comparison of performance against clinical quality indicators for acute medicine using patient level data is published elsewhere.(11–14)

The organisational survey collected data detailing hospital size (including number of beds on the AMU and total number of inpatient beds); in addition, SAMBA22 contained more detailed questions regarding medical SDEC services, designed to assess adherence to national recommendations and describe service availability (appendix 1).(15) Units were asked regarding provision of condition-specific ambulatory pathways; these were selected from guidance regarding ambulatory care sensitive conditions,(16) the AEC directory,(4) and service evaluations presented at SAM conferences over the preceding 12 months.

The terminology used to refer to SDEC changed across the time period covered by this dataset. AEC and SDEC services as they relate to medical patients are assumed to be equivalent, and the terms have been used interchangeably across the data collection period.

Patient level data was combined from SAMBA19 (20^th^ June 2019), Winter SAMBA20 (23^rd^ January 2020), SAMBA21 (June 2021), SAMBA22 (17^th^ June 2022) and SAMBA23 (22^nd^ June 2023). Data was aggregated to allow comparison where levels of measurement had changed over time, for example increased granularity recorded within the “time to consultant review” measure in more recent data collection.

All patient-level data submitted to SAMBA is anonymised. Patient-level data was divided into planned and unplanned attendances. Age (in bands) and gender is recorded for both planned and unplanned attendances. Reason for attendance was available for planned attendances in 2022/23. For unplanned attendances, the location of initial clinical assessment and first assessment by the medical team was recorded, categorised as Emergency Department (ED), AMU, SDEC or other locations. Records where neither the location of initial assessment nor medical assessment was documented were excluded from analysis (63 patients, 0.2% of unplanned admissions in the dataset). Patients presenting to non-standard units (frailty units, specialist cancer centres, and stand-alone SDEC services) were excluded from analysis as patient pathways into and through these services is likely to be different from standard acute medicine services.(17)

Data was analysed using Microsoft Excel and STATA 16/18 (StataCorp LP, College Station, TX, USA). Descriptive statistics were used to summarise results, with group comparisons made using Chi square test, or Fisher’s exact test where expected cell counts were <5. For group comparisons of continuous variables, Kruskal-Wallis and Mann Whitney U tests were used for data that was not normally distributed. For analysis of survey data describing SDEC services included only in SAMBA22, participating units were stratified based on hospital size, into three groups: smaller (<400 inpatient beds, 41 hospitals), medium (400-599 inpatient beds, 50 hospitals) and larger (≥600 inpatient beds, 52 hospitals). Where questions regarding organisational structure were asked in only selected rounds of data collection, any associated comparison to patient-level data is limited to those years. Correlation between variables that were not normally distributed was assessed using Spearman’s rank correlation. Comparison between attendance numbers and SDEC and AMU size was performed using data from SAMBA23 only. Logistic regression using backwards selection was used to assess factors affecting likelihood of meeting the same-day discharge target (one third of admissions discharged without inpatient admission, outlined in NHS Long Term Plan)(7) using data from units that participated in all rounds of data collection 2021-2023; odds ratios and confidence intervals are reported. A p value of <0.05 was considered statistically significant throughout.

## Results

### Organisational survey

Of the 149 UK hospitals that participated in SAMBA22, 140 hospitals responded to the organisational questions regarding SDEC; this included 122 hospitals from England (87% of participating units), 8 from Scotland (6%), 6 from Wales (4%) and 4 from Northern Ireland (3%)(Supplementary Table 1). Hospital size ranged from 0-1700 inpatient beds (median 520, IQR 369-688), with one stand-alone SDEC service at a site with no inpatient beds. Only one hospital that responded did not have a medical SDEC service. SDEC services were physically separate from the AMU in 89% (120 units).

SDEC units were open for a median of 12 hours (IQR 11-14 hours, range 4-24 hours) although nine SDEC units (6.5%) were open 24 hours a day.

#### Recommended standards

Comparison to recommended standards for SDEC are shown in Supplementary Table 2. Overall, a consultant was physically available throughout SDEC opening hours in only 64% (89 units). There was no difference in the proportion of unplanned attendances assessed in SDEC service comparing those with a consultant available to those without (Mann Whitney U, p=0.867), however when comparing SDEC discharge rates, more units that had a consultant available discharged over 80% of patients assessed in SDEC the same day (72% vs 45%, Chi square p=0.007).

A nominated clinician had overall leadership of SDEC in 85% (118 units); of these, 87% (103) were consultant physicians, 3% (4) were nurses, 6% (7) were advanced clinical practitioners (ACPs) and 3% (4) were specialist or specialty doctors. Compliance with standards for SDEC did not vary with hospital size, including availability of private consultation areas, patient feedback collection and contact with non-attenders.

Overall, 113 SDEC units (81%) had a Standard Operating Procedure (SOP), 17 (12%) did not and 9 (7%) were unsure; this did not vary with hospital size (p=0.51).

#### Pathways into SDEC services

##### Triage

Eighty-five hospitals (61%) used a specific scoring system to assist in identifying patients suitable for treatment in SDEC; 42% (58 units) used NEWS2(18), 11% (15 units) used centre-specific criteria, 6% (8) used the Amb score,(19) 2% (3) used the Glasgow Admission Prediction Score (GAPS)(20), and one unit (0.7%) used Clinical Frailty Scale (CFS).(21)

There was no difference in the proportion of unplanned attendances assessed in SDEC comparing those that used a scoring system and those that did not (Mann Whitney U, p=0.075), and no difference in the proportion of patients assessed in SDEC who were discharged the same day when comparing units that did not use a criteria to units using NEWS, or an Amb or GAPS score (Kruskal Wallis, p=0.162).

Thirty-four services (24%) exclusively saw patients attending with specific conditions (e.g. deep vein thrombosis (DVT)) or on protocolised pathways; of these, 28 (82%) had a SOP. Comparing these services to those that were not limited to specific conditions/pathways, there was no difference in the proportion of unplanned admissions seen in SDEC (Mann Whitney U, p=0.247), the proportion of all unplanned attendances discharged without overnight admission (p=0.593), or the proportion of patients seen in SDEC discharged without overnight admission (p=0.157).

Thirty-two units (23%) did not accept patients that required assistance with mobility, 16 units (12%) did not accept patients confined to a chair, and 104 units (75%) did not accept patients confined to a bed.

##### Referral

Accepted referral sources are shown in Table 1. Most units (82%) accepted patients to SDEC without requiring prior full clinical review; of these, 98 (86%) had a SOP. Most units (51%, 71 units) accepted referrals from the ED without discussion with the medical team. Of the 34 units accepting only selected conditions/pathways, 82% (24 units) accepted these patients from ED without assessment by an Emergency Medicine (EM) clinician.

**Table 1:** Form of referral accepted by SDEC units. ED: Emergency Department; GP: General Practice.

| Pathways accepted for referral to SDEC | Overall (N = 139) |  | Smaller hospitals (N = 40) |  | Medium hospitals (N = 47) |  | Larger hospitals (N = 52) |  | P value |
| --- | --- | --- | --- | --- | --- | --- | --- | --- | --- |
|  | % | n | % | n | % | n | % | n |  |
| Referral from ED triage without full clinician review | 82 | 114 | 83 | 33 | 81 | 38 | 83 | 43 | 0.97 |
| Referral from ED without discussion with medical team | 51 | 71 | 45 | 18 | 55 | 26 | 52 | 27 | 0.62 |
| Referral from GP without further clinician review | 88 | 122 | 88 | 35 | 89 | 42 | 87 | 45 | 0.91 |
| Direct referral from paramedics | 63 | 87 | 70 | 28 | 62 | 29 | 58 | 30 | 0.48 |

#### SDEC services available

##### Specific conditions

Of the pre-specified conditions included, condition-specific ambulatory pathways were most common for pulmonary embolism (PE) and DVT, both overall and when stratified by hospital size (Table 2). Nine SDEC units (6%) had no condition-specific ambulatory pathways available, and an additional three units (2.1%) did not report provision of any ambulatory pathways for the conditions listed. Provision of condition-specific pathways was similar when stratifying by hospital size; larger hospitals were more likely to have guidelines for ambulatory management of papilledema.

**Table 2:** The provision of condition-specific ambulatory pathways in SDEC units. Preselected conditions, ordered by number of units with ambulatory pathway. *Fishers exact test

| Ambulatory pathway conditions | Overall (N = 139) |  | Small hospitals (N = 40) |  | Medium hospitals (N = 47) |  | Large hospitals (N = 52) |  | P value |
| --- | --- | --- | --- | --- | --- | --- | --- | --- | --- |
|  | % | n | % | n | % | n | % | n |  |
| Deep vein thrombosis | 86 | 119 | 83 | 33 | 79 | 37 | 94 | 49 | 0.07 |
| Pulmonary embolism | 83 | 116 | 80 | 32 | 79 | 37 | 90 | 47 | 0.23 |
| Chest Pain | 58 | 80 | 58 | 23 | 60 | 28 | 56 | 29 | 0.93 |
| Cellulitis | 55 | 77 | 53 | 21 | 55 | 26 | 58 | 30 | 0.88 |
| Anaemia | 51 | 71 | 43 | 17 | 47 | 22 | 62 | 32 | 0.15 |
| Headache | 47 | 66 | 38 | 15 | 45 | 21 | 58 | 30 | 0.14 |
| Atrial fibrillation | 42 | 58 | 38 | 15 | 43 | 20 | 44 | 23 | 0.80 |
| Acute kidney injury | 29 | 40 | 23 | 9 | 32 | 15 | 31 | 16 | 0.58 |
| Deranged Liver function tests | 27 | 38 | 25 | 10 | 28 | 13 | 29 | 15 | 0.92 |
| Heart failure | 27 | 37 | 25 | 10 | 21 | 10 | 33 | 17 | 0.42 |
| Papilledema | 25 | 35 | 23 | 9 | 15 | 7 | 37 | 19 | 0.04 |
| COVID-19 | 15 | 21 | 8 | 3 | 13 | 6 | 23 | 12 | 0.10 |
| None | 6 | 9 | 10 | 4 | 6 | 3 | 4 | 2 | 0.52* |

##### Planned reattendance

Ninety-nine percent of units (136/138) enabled booked patients to return to SDEC, utilised by acute medicine (98.5%, 134 units), EM (79%, 108 units) and inpatient medical wards (63%, 86 units). Returning patients were booked to timeslots in 72% of units (97/135). Eighty-two percent of those offering booked return (111/136) had an SDEC SOP.

Services offered through planned SDEC reattendance varied (Table 3). Specialty review was the least common service, offered in 72% (100 units).

**Table 3:** Services available for planned patients returning to SDEC. IV: intravenous; CT: computed tomography; US: ultrasound; DVT: deep vein thrombosis. *Fishers exact test

|  | Overall<br>(N=139) |  | Small hospitals<br>(N=40) |  | Medium hospitals<br>(N=47) |  | Larger hospitals<br>(N=52) |  | P value |
| --- | --- | --- | --- | --- | --- | --- | --- | --- | --- |
|  | % | *n* | % | *n* | % | *n* | % | *n* |  |
| Acute medicine review | 96 | 134 | 100 | 40 | 98 | 46 | 92 | 48 | 0.19* |
| IV antibiotics | 89 | 124 | 85 | 34 | 89 | 42 | 92 | 48 | 0.53 |
| CT scans | 98 | 136 | 98 | 39 | 98 | 46 | 98 | 51 | 1.0* |
| US scans | 98 | 136 | 98 | 39 | 96 | 45 | 100 | 52 | 0.39* |
| Anaemia treatment | 91 | 127 | 95 | 38 | 85 | 40 | 94 | 49 | 0.19* |
| DVT rule out | 88 | 123 | 88 | 35 | 85 | 40 | 92 | 48 | 0.52 |
| Specialty review | 72 | 100 | 60 | 24 | 79 | 37 | 75 | 39 | 0.13 |

#### Patient level data

Patient level data was available for 39,722 patients, of which 4,342 (11%) were planned reattendances to SDEC services. Excluding patients presenting to non-standard units and those without assessment location data available, data for 34,948 unplanned attendances from 188 units were available for analysis (Supplementary Figure 1). Distribution by UK nation is shown in Supplementary Table 1.

##### Unplanned attendances

The proportion of unplanned admissions receiving their initial medical team assessment in SDEC has increased over time (Figure 1), with 29.8% of unplanned attendances in SAMBA23 assessed in SDEC services (Supplementary Table 3, Chi Square p<0.005).

**Figure 1:**
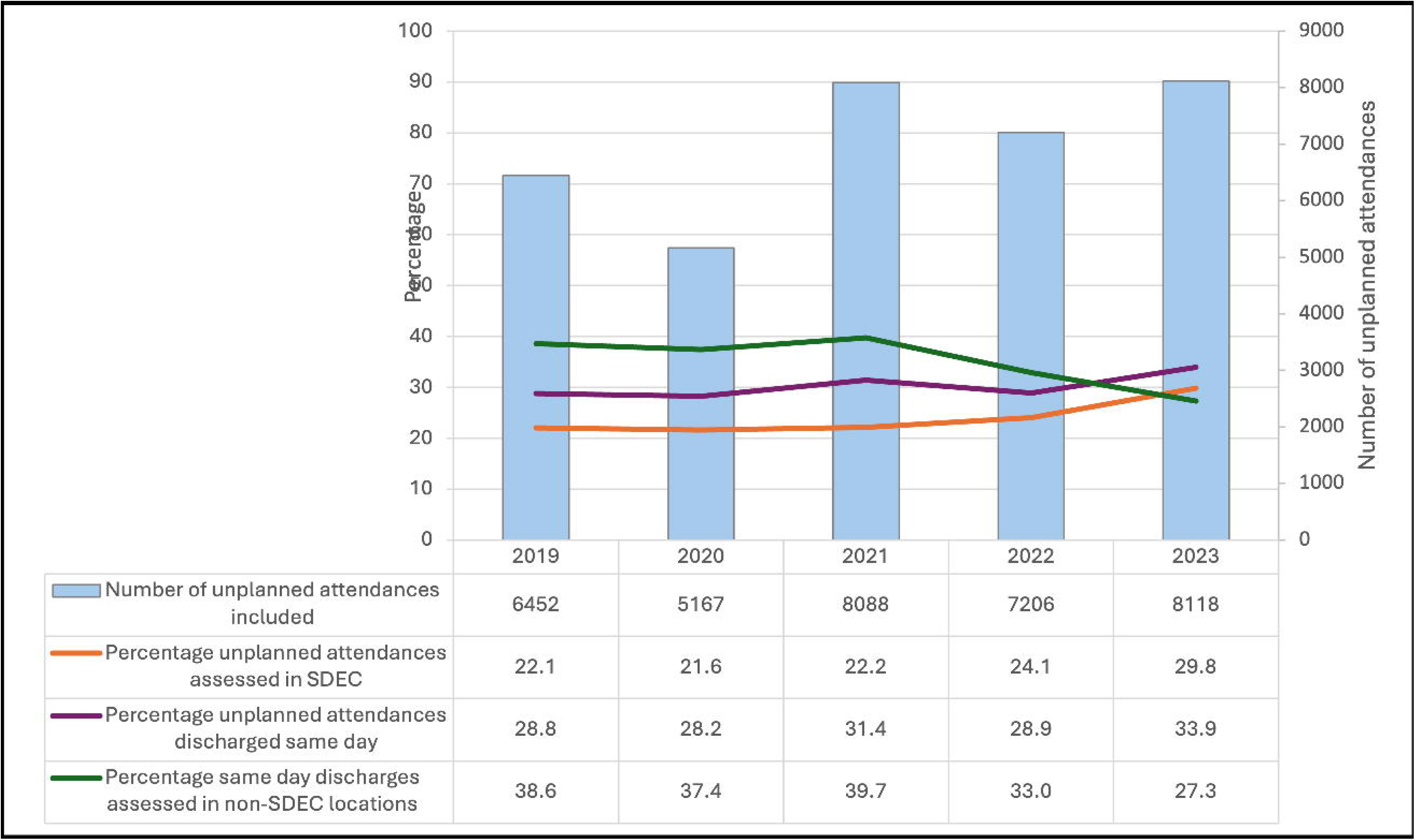
Comparison of unplanned attendances and same day discharges by year. SDEC: Same Day Emergency Care.

The proportion of patients receiving medical assessment in SDEC services varied between hospitals (Figure 2a, median 21.8%, IQR 12.5-31.7%, range 0-71.1%).

**Figure 2:**
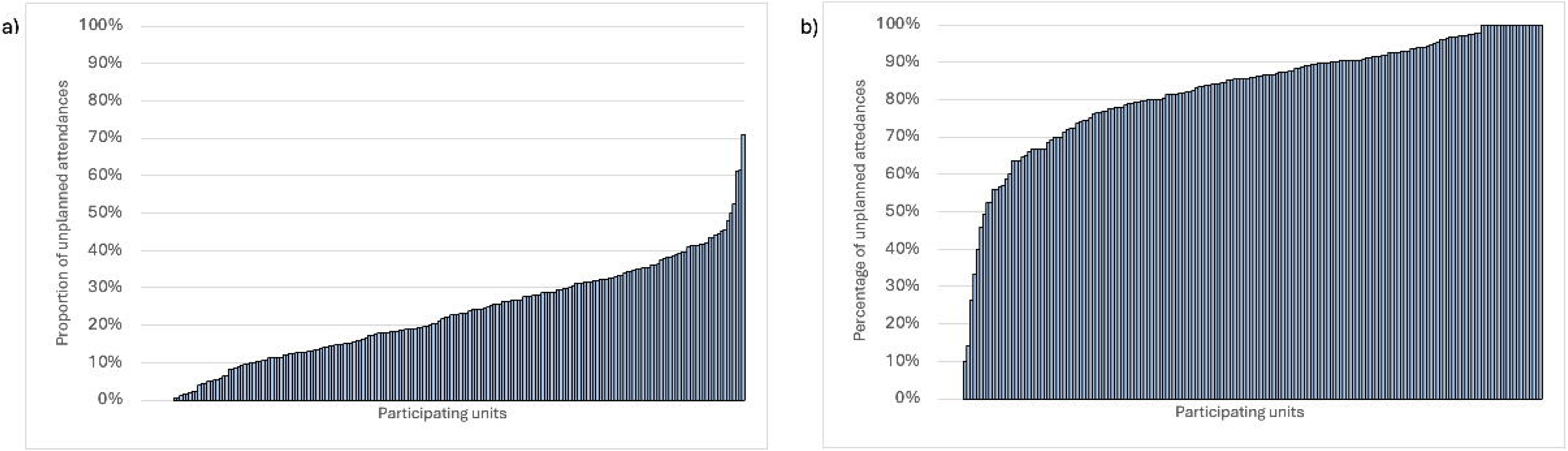
Comparison of performance by site. 2a: Proportion of unplanned attendances receiving their medical assessment within Same Day Emergency Care services by site. Units ordered along x-axis by proportion assessed within SDEC. Median unit performance: 21.8%. 2b: Proportion of patients receiving medical assessment in SDEC who were discharged same day, by unit. Units ranked along x-axis by performance.

A higher proportion of patients in England (25.5%) were assessed within SDEC (or their counterpart) services than in other UK nations (Scotland 10.2%, Wales 15.5%, Northern Ireland 6.7%, p<0.005).

Amongst English sites, units assessed a median of 24.5% of patients in SDEC (IQR 14.6-32.6%, range 0.0-71.1%).

Overall, 17.6% of patients receiving their medical assessment in SDEC had been assessed in another location prior to this; this was lower in 2023 (14.6%) compared to previous years (2019: 19.3%, 2020: 17.4%, 2021: 18.9%, 2022: 19.4%, Chi square <0.001).

##### Patient characteristics by assessment location

Patient demographics and acuity (assessed by NEWS score) were compared for patients receiving medical assessment in SDEC compared to those assessed in the ED, AMU and other locations (Table 4). A higher proportion of female patients received medical assessment in SDEC compared to male patients (26.3% vs 22%, p<0.005). A lower proportion of those assessed in SDEC were aged ≥70 years compared to those assessed in other locations (30% of patients assessed in SDEC, 54% average across other locations, Chi square <0.005). This also equates to a lower proportion of patients aged over 70 receiving their initial assessment in SDEC compared to those aged under 70 (15.1% vs 32.8%).

**Table 4:** Patient characteristics by location of first assessment by the medical team. Data for 34,897 unplanned attendances. SDEC: Same Day Emergency Care; ED: Emergency Department; AMU: Acute Medical Unit; NEWS: National Early Warning Score; CFS: Clinical Frailty Score. *ANOVA.

|  | SDEC (n= 8466) |  | ED (n= 15639) |  | AMU (n=9745) |  | Other (n=1047) |  |  |
| --- | --- | --- | --- | --- | --- | --- | --- | --- | --- |
| Patient population | N | % | N | % | N | % | N | % | P value |
| Age (years) |  |  |  |  |  |  |  |  | <0.005* (KW) |
| 16-29 | 1015 | 12.0% | 1003 | 6.4% | 637 | 6.5% | 82 | 7.8% |  |
| 30-39 | 1063 | 12.6% | 963 | 6.2% | 649 | 6.7% | 72 | 6.9% |  |
| 40-49 | 1150 | 13.6% | 1096 | 7.0% | 745 | 7.6% | 72 | 6.9% |  |
| 50-59 | 1375 | 16.2% | 1794 | 11.5% | 1040 | 10.7% | 103 | 9.8% |  |
| 60-69 | 1318 | 15.6% | 2225 | 14.2% | 1502 | 15.4% | 131 | 12.5% |  |
| 70-79 | 1414 | 16.7% | 3420 | 21.9% | 2183 | 22.4% | 198 | 18.9% |  |
| 80-89 | 951 | 11.2% | 3798 | 24.3% | 2201 | 22.6% | 282 | 26.9% |  |
| 90+ | 180 | 2.1% | 1340 | 8.6% | 788 | 8.1% | 107 | 8.6% |  |
| >70 years | 2545 | 30.1% | 8558 | 54.7% | 5172 | 53.1% | 587 | 56.1% | <0.005 |
| Gender |  |  |  |  |  |  |  |  |  |
| Female | 4860 | 57.4% | 7953 | 50.9% | 5086 | 52.2% | 577 | 55.1% | <0.005 |
| Care home resident<br>(missing=36) | 81 | 1.0% | 1181 | 7.6% | 661 | 6.8% | 66 | 6.3% | <0.005 |
| Discharged from hospital in preceding 30<br>days (missing=18) | 1132 | 13.4% | 3246 | 20.8% | 1983 | 20.4% | 229 | 21.9% | <0.005 |
| Arrival to hospital* (missing=2) |  |  |  |  |  |  |  |  | 0.04* (KW) |
| 00:00-08:00 | 377 | 4.5% | 2918 | 18.7% | 1637 | 16.8% | 156 | 14.9% |  |
| 08:00-20:00 | 7947 | 93.9% | 10449 | 66.8% | 6653 | 68.3% | 7947 | 72.7% |  |
| 20:00-23:59 | 142 | 1.7% | 2271 | 14.5% | 1454 | 14.9% | 142 | 12.4% |  |
| NEWS* (missing=744) |  |  |  |  |  |  |  |  | <0.005* |
| 0 | 4627 | 56.8% | 4352 | 28.4% | 3099 | 32.1% | 432 | 41.9% |  |
| 1 | 2111 | 25.9% | 2999 | 19.6% | 2102 | 21.8% | 221 | 21.4% |  |
| 2 | 787 | 9.7% | 2089 | 13.6% | 1429 | 14.8% | 131 | 12.7% |  |
| 3 | 358 | 4.4% | 1694 | 11.1% | 1072 | 11.1% | 95 | 9.2% |  |
| 4 | 147 | 1.8% | 1170 | 7.6% | 683 | 7.1% | 43 | 4.2% |  |
| 5 | 61 | 0.7% | 880 | 5.7% | 481 | 5.0% | 32 | 3.1% |  |
| 6 | 36 | 0.4% | 736 | 4.8% | 309 | 3.2% | 26 | 2.5% |  |
| 7+ | 20 | 0.2% | 1397 | 9.1% | 482 | 5.0% | 52 | 5.0% |  |
| Missing | 319 |  | 322 |  | 88 |  | 15 |  |  |
| NEWS ≥3 | 622 | 7.6% | 5877 | 38.4% | 3027 | 31.4% | 248 | 24.0% | <0.005 |
| CFS (in over 70s) (n=11957) | (1720) |  | (6399) |  | (3429) |  | (409) |  | <0.005 |
| 1 | 166 | 9.7% | 256 | 4.0% | 170 | 5.0% | 22 | 5.4% |  |
| 2 | 283 | 16.5% | 466 | 7.3% | 265 | 7.7% | 23 | 5.6% |  |
| 3 | 478 | 27.8% | 1014 | 15.8% | 579 | 16.9% | 61 | 14.9% |  |
| 4 | 349 | 20.3% | 1122 | 17.5% | 627 | 18.3% | 64 | 15.6% |  |
| 5 | 206 | 12.0% | 1116 | 17.4% | 564 | 16.4% | 70 | 17.1% |  |
| 6 | 153 | 8.9% | 1211 | 18.9% | 666 | 19.4% | 87 | 21.3% |  |
| 7 | 66 | 3.8% | 877 | 13.7% | 413 | 12.0% | 58 | 14.2% |  |
| 8 | 13 | 0.8% | 249 | 3.9% | 102 | 3.0% | 18 | 4.4% |  |
| 9 | 6 | 0.3% | 88 | 1.4% | 43 | 1.3% | 6 | 1.5% |  |
| CFS ≥5 | 444 | 25.8% | 3541 | 55.3% | 1788 | 52.1% | 239 | 58.4% | <0.005 |

Care home residents were less likely to have received initial medical assessment in SDEC than patients living at home or in other settings (4.1% vs 25.5%, p<0.005). In those aged over 70 years, 7.4% of patients with CFS ≥5 were seen in SDEC, compared to 30.4% of those with a CFS 1-4 (p<0.005). Patients who had been in hospital in the preceding 30 days were also less likely to have been assessed in SDEC (17.2% vs 25.9%, p<0.005).

Only 7.6% of patients seen within SDEC services had a NEWS score of ≥3 on arrival to hospital, compared to 38.4% of those assessed in the Emergency Department and 31.4% of those assessed on AMU (Chi square p<0.005).

##### Patient characteristics over time

Patient demographics and acuity in patients receiving medical assessment within SDEC services were compared across data collection periods (Supplementary Table 4); there was no significant change in the proportion of patients seen within SDEC services that were aged over 70 years, female or care home residents.

There was no significant difference in the NEWS scores of patients seen within SDEC services when comparing the summer data collection periods - a higher proportion of patients assessed in SDEC services in the January 2020 cohort had a NEWS score ≥3 compared to the summer cohorts (p=0.01).

Adjusting for time of arrival and source of referral, patients who were care home residents, male, or had been recently discharged from hospital were less likely to have been assessed within SDEC services (Table 5). Odds of assessment within SDEC services reduced with increasing age (over the age of 50) and increasing NEWS score. The likelihood of assessment in SDEC was higher in recent time periods.

**Table 5:** Logistic regression model for likelihood of medical assessment within SDEC services. Pseudo R ^2^ = 0.272. NEWS: National Early Warning Score.

|  | OR | P value | 95% CI |
| --- | --- | --- | --- |
| Age |  |  |  |
| 16-29 | *Reference* |  |  |
| 30-39 | 0.99 | 0.863 | 0.87-1.12 |
| 40-49 | 0.94 | 0.308 | 0.82-1.06 |
| 50-59 | 0.73 | <0.005 | 0.65-0.82 |
| 60-69 | 0.55 | <0.005 | 0.49-0.62 |
| 70-79 | 0.39 | <0.005 | 0.34-0.43 |
| 80-89 | 0.27 | <0.005 | 0.24-0.30 |
| 90+ | 0.16 | <0.005 | 0.14-0.20 |
| Gender |  |  |  |
| Female | *Reference* |  |  |
| Male | 0.83 | <0.005 | 0.78-0.88 |
| Other | 0.89 | 0.887 | 0.18-4.44 |
| Care home residence |  |  |  |
| No | *Reference* |  |  |
| Yes | 0.30 | <0.005 | 0.23-0.38 |
| Recent discharge |  |  |  |
| No | *Reference* |  |  |
| Yes | 0.70 | <0.005 | 0.64-0.76 |
| NEWS score | 0.64 | <0.005 | 0.62-0.65 |
| Time of arrival |  |  |  |
| 00:00-07:59 | 0.27 | <0.005 | 0.24-0.30 |
| 08:00-19:59 | *Reference* |  |  |
| 20:00-23:59 | 0.11 | <0.005 | 0.09-0.13 |
| Source of referral |  |  |  |
| ED | *Reference* |  |  |
| 111 | 1.12 | 0.569 | 0.76-1.66 |
| GP | 4.17 | <0.005 | 3.90-4.45 |
| Other hospital | 1.36 | 0.026 | 1.04-4.45 |
| Own hospital (OPD) | 3.89 | <0.005 | 3.31-4.58 |
| Own hospital (other) | 3.36 | <0.005 | 2.82-4.00 |
| Paramedic | 1.00 | 0.96 | 0.82-1.23 |
| Year |  |  |  |
| 2019 | *Reference* |  |  |
| 2020 | 1.12 | 0.039 | 1.00-1.24 |
| 2021 | 1.17 | 0.001 | 1.07-1.28 |
| 2022 | 1.39 | <0.005 | 1.27-1.53 |
| 2023 | 1.75 | <0.005 | 1.60-1.92 |

#### Outcomes

Outcome data within 7 days was available for 34,621 patients. Six patients (0.1%) receiving medical assessment in SDEC died within 7 days.

##### Same day discharge

Overall, 30.6% of all patients referred to AIM were discharged without overnight admission; 82.4% of patients receiving medical assessment in SDEC were discharged without overnight admission (Supplementary Table 5). The proportion of patients discharged without overnight admission was highest in 2023 (Chi square <0.001). Likelihood of same-day discharge was associated with patient factors similar to those influencing likelihood of assessment within SDEC services (Supplementary Table 6).

The likelihood of meeting the NHS target for one third of patients being discharged same day was assessed for units participating in all rounds of 2021-2023. The odds of a unit meeting the target of one third increased with increasing number of unplanned admissions and an increased percentage of patients that were daytime arrivals or were GP referrals, and decreased with increasing percentage of patients that were aged over 70 or had a NEWS2 ≥3 (Table 6).

**Table 6:**
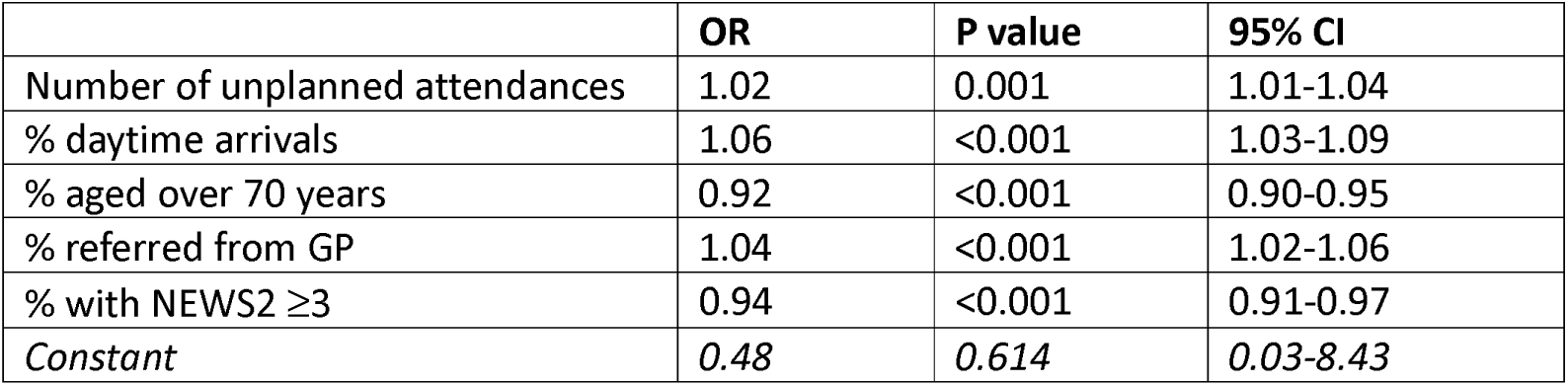
Logistic regression assessing likelihood of unit meeting target for same day discharges. Target: one third of unplanned attendances discharged without inpatient admission. Pseudo R ^2^ = 0.2838. Analysis limited to units participating in three consecutive years (2021–2023); 115 units with 17,159 unplanned medical attendances. OR: odds ratio; CI: confidence interval; GP: general practice; NEWS: National Early Warning Score

3672 patients discharged same day received their medical assessment in non-SDEC locations; this was 34.7% of those discharged without overnight admission, and 10.6% of patients overall. This has decreased over time (Figure 1; 2019: 38.6%, 2020: 37.4%, 2021: 39.7%, 2022: 33.0%, 2023: 27.3%, Chi square <0.001).

##### SDEC discharge rates

The proportion of unplanned attendances assessed in SDEC who were discharged same day varied between units (Figure 2b, median 85.7%, IQR 74.9-92.59, range 0-100%). Comparison by time period is shown in Figure 1 & Supplementary Table 7. In the most recent data collection, the conversion rate to inpatient admission was <20% in most units (69.3%, 97 units), 20-30% in 12.1% (17 units) and >30% in 18.6% (26 units).

Data regarding unplanned re-attendances within 7 days was available for 6625 unplanned attendances who received medical assessment in SDEC and were discharged same day; 4.7% (309 patients) had an unplanned reattendance to ED (148 patients), AMU (23 patients) and/or SDEC (137 patients).

##### Planned reattendances

Of the 4,342 patients included as a planned reattendance, reason for this reattendance was recorded for 2,450 (SAMBA22 and SAMBA23 only); 35.6% were ≥70 years, and 55.7% were female. Clinical review was the most common reason for reattendance (1163 patients, 48%), followed by repeat blood tests (642, 26%), DVT investigation/treatment (456, 19%), imaging (432, 18%), intravenous (IV) antibiotics (209, 9%), non-antibiotic IV medication (201, 8%), and ambulatory PE diagnosis/treatment (143, 6%). Ninety percent were assessed by a clinician during this reattendance (2201/2445), with initial assessment by doctor more junior than registrar in 32% (686/2182), registrar in 26% (566), ANP/ACP in 24% (515), consultant in 13% (280) and PA in 3% (56).

92.6% of planned reattendances were discharged home without overnight admission (2229/2407), 3.6% were discharged within the next 7 days, 1% (25) discharged against medical advice, and 1 patient (0.04%) died in hospital within 7 days.

##### Attendances and SDEC size

Number of planned and unplanned attendances seen within SDEC services was compared to hospital size, AMU size and number of assessment spaces on AMU using data from SAMBA23. For units with this data available, the number of unplanned attendances receiving their medical assessment in SDEC ranged from 0-61 (median 14, IQR 6-21), with 16 units (11.3%) reporting that no patients received their medical assessment in SDEC on SAMBA day. The number of planned reattendances seen in SDEC ranged from 0-52 (median 7, IQR 3-12), with 18 units (12.7%) reporting no planned reattenders on the day of data collection. The proportion of patients seen within SDEC that were planned returners ranged from 0-100% (median 34.9%, IQR 19.6-51.4%).

There was a moderate positive correlation between the number of planned reattenders and the number of unplanned attendances receiving medical review in SDEC (r=0.46, p<0.005), and between the number of unplanned admissions and the proportion of unplanned admissions assessed within SDEC (r=0.42, p<0.005).

SDEC services had a median number of ten assessment spaces (IQR 6-13). There was no significant correlation between the number of SDEC assessment spaces and size of hospital (assessed by number of inpatient beds, r=0.12, p=0.16) or number of beds on the AMU (r=0.07, p=0.38), however there was a moderate correlation between number of AMU beds and hospital size (r=0.51, p<0.005).

Although there was a moderate correlation between the number of unplanned attendances and both hospital size (r=0.59, p<0.005) and number of beds on AMU (r=0.45, p<0.005), the number of SDEC assessment spaces available was very weakly correlated with the total number of unplanned attendances (r=0.17, p=0.046), and weakly correlated with the number of unplanned attendances receiving medical assessment in SDEC (r=0.21, p=0.013). There was no significant association between the number of planned reattenders and number of SDEC assessment spaces (r=0.13, p=0.14), AMU beds (r=0.19, p=0.02) or inpatient beds (r=0.18, p=0.03).

Opening hours and number of assessment spaces were used to calculate the total space available daily (136 units with available data). The number of patients that would require assessment within SDEC to achieve the recommended discharge rate of one third was calculated from the total number of unplanned admissions seen, assuming a 20% conversion rate from SDEC assessment. Each patient was assumed to require one hour of assessment space. Two units (1.5%) did not have enough space available to assess the required patient numbers.

## Discussion

Medical SDEC services are a key component of assessment pathways for acute medical patients within the UK, however there is considerable variation in provision of these services, in terms of both operational factors and clinical delivery.

The proportion of medical patients assessed through SDEC services has increased over the last five years, but considerable variation remains between hospitals, and between UK nations. Although NHS England has consistently advocated for increased use of SDEC,(6,7,22,23) and NHS Scotland has supported its counterpart Rapid Assessment and Discharge, there has arguably been slower adoption in other Wales and Northern Ireland.(24) This may have influenced the higher proportion of patients seen in SDEC services within English hospitals in this study.

Whether an individual hospital met the ‘Long Term Plan’ target(7) that one third of patients be discharged without overnight admission was influenced by the patient population, including the proportion of patients aged over 70 and with high acuity (assessed by NEWS2 score). Hospitals with more unplanned medical attendances were more likely to achieve the target; increased attendances have prompted higher SDEC activity to maintain patient flow and mitigate service pressures. The factors identified here as relating to achievement of target discharge rate are not amenable to alteration by hospitals, but may provide an understanding of why individual services may not achieve the target, and support approaches targeting improved delivery of SDEC and provision of community services for older adults.

Most SDEC services (69%) had a conversion rate to inpatient admission of less than 20%; only 12% of units had a conversion rate of 20-30% as recommended in the SAMEDAY strategy.(23) There is little evidence available assessing the optimum SDEC conversion rate; accepting patients into SDEC services with a higher chance of admission may impact ability to deliver assessment to low risk patients due to the limited space available, while some patients discharged without admission may have been suitable for assessment through other care pathways, such as primary care, community or outpatient services. Challenges in access to these services, perceived or genuine, may contribute to the increase in patients seen through SDEC services; greater understanding of these factors is likely to be beneficial to UEC services but is beyond the scope of this study.

Our results suggest there is low mortality in patients currently assessed through SDEC services and discharged without overnight admission (0.1% within 7 days), with 5% of patients discharged after SDEC assessment having an unplanned reattendance within 7 days. The methodology used does not allow further exploration of mortality cases, and data could not be linked to external sources to confirm mortality or 30-day readmission rates.

Services had an average of 10 spaces available for patient assessment within SDEC; although the size of a hospital’s AMU had some correlation to hospital size and the number of unplanned admissions seen daily, there was only weak association between these factors and the space available for SDEC suggesting the SDEC footprint was not mapped against potential demand. Only 1.5% of units were estimated to lack the necessary space to assess the patient numbers required to meet the current discharge targets, however this assumes availability of staffing, and uniform flow of patients into the service. There is currently no guidance describing how to calculate the space needed to deliver SDEC effectively based on expected or desired patient flow, and greater understanding of optimal physical set-up, including size, is needed. Our results suggest greater physical space alone currently does not equate to increased clinical activity. There was some association between the number of planned and unplanned attendances seen within SDEC, suggesting services seeing more unplanned attendances also provide more scheduled care delivery; while this may reflect greater performance driving increases in both, it may be that increased levels of scheduled care are required to facilitate delivery of unscheduled care through SDEC.(25)

It is likely that some patients suitable for SDEC are still receiving medical assessment in other locations: almost 30% of patients discharged without overnight admission were assessed in non-SDEC locations. Many of these may have been suitable for SDEC, however a zero-day length of stay should not be assumed to equate to suitability for SDEC. Correct identification of suitable patients is vital for effective delivery of SDEC.(18) Over 80% of SDECs accept patients from ED triage without full clinician review, necessitating a robust process to ensure that only patients likely to be discharged after medical team intervention are directed through SDEC services. Inappropriate identification of patients could result in delayed delivery of care to those requiring inpatient care or more appropriately managed through other pathways, as well as reducing the ability to deliver SDEC to those who are suitable. An understanding of how these processes can be operationalised and delivered effectively and the impact on patient outcomes is needed.

Over a third of SDEC units did not use a screening tool to identify suitable patients. This may make patient selection more subjective, however use of a screening tool was not associated with higher rates of SDEC assessment or same day discharge in our results. NEWS2, which identifies patients at high risk of impending deterioration, was the most common tool used to identify suitable patients, although cut-offs used were not recorded.(18,26) NHS Improvement suggests only those with NEWS <4 be considered for SDEC due to potential clinical instability, but not all deterioration is preceded by a raised NEWS, and use without consideration of other factors may not prevent inappropriate referrals.(2,27) Scores incorporating additional features have been suggested,(28) including the Amb score and Glasgow Admission Prediction Score (GAPS),(19,20) however their discriminatory ability appears to be lower when applied outside the original setting.(29–31)

Patient factors such as age and recent hospital attendance, that feature in these scoring systems, were associated with decreased likelihood of assessment within SDEC services, and of same day discharge, within our analysis. This suggests these features are linked to suitability for SDEC, although this may reflect availability of services that facilitate discharge in selected patient groups. Although the likelihood of receiving assessment within SDEC was influenced by the patient factors discussed here, other features not included within the patient-level data may play a role. There are likely to be significant barriers to medical SDEC services for those with reduced mobility; a quarter of units did not accept patients requiring assistance with mobility and 1 in 10 did not accept patients confined to a chair. Our results suggest that older patients were less likely to be assessed through SDEC services, and less likely to be discharged without inpatient admission; previous analysis suggested assessment in SDEC was less common in those with frailty or presenting with a geriatric syndrome.(32) Although this may disproportionately impact older adults and those with frailty, these patients may now be supported by the increased emphasis on frailty SDEC services.(17,33) How these services are delivered and interact with medical SDEC, including the clinical conditions amenable to management through these services, requires further evaluation.

Condition-specific pathways can improve patient outcomes and reduce cost, with multiple acute medical conditions suggested as suitable for management through SDEC.(34,35) Despite this, aside from DVT and PE, many SDEC units lacked condition-specific ambulatory pathways. While this risks inconsistent service provision within and between SDEC units, there is limited evidence regarding how condition-specific SDEC pathways may impact quality of care, delays in management, resource utilisation, and patient experience.

Adherence to SDEC standards recommended by SAM and RCPE is variable.(15) More than a third of participating services did not have a consultant physically available throughout operational hours. This may impact care delivery by introducing delays or inefficiencies when junior clinical staff require input in complex cases; our results suggest units with consultant presence were more likely to discharge a high proportion (>80%) of the patients assessed in SDEC. However, the SAMEDAY strategy now recommends a more lenient target, suggesting “access to an appropriate consultant” as a minimum requirement;(23) how the recommended standards can be delivered in practice, and barriers such as workforce limitations, should be explored.

Although there are recommended standards, the operational protocols and pathways used within SDEC units remain the responsibility of local teams, allowing services to be tailored to the requirements of local pressures and populations. Standard operational policies are recommended to ensure SDEC is not used for patients that would be more appropriately managed through alternative pathways or inpatient care;(36) 12% of units did not have a SOP, and an additional 7% were unsure. Similar figures were seen in services accepting ED triage referrals without full clinician review, and services allowing booked patient reattendance, where variation may increase risk and a more structured approach may be beneficial. There is concern nationally that SDEC services may be used to house patients that have spent prolonged periods in the ED prior to transfer, including those awaiting diagnostic test results and where medical physician input is not required,(25,36) however evaluation of this was beyond the scope of this study.

Across the 48 hours of SAMBA22 and SAMBA23, almost 2500 patients were seen in SDEC as a planned reattendance, most commonly for clinical review. Despite the large numbers seen through this route, there is limited guidance describing how planned reattendance to SDEC services should be used to facilitate discharge outside of specific conditions, such as PE,(37) and little evidence evaluating how these attendances impact patient care, acute medicine resource use and service pressures.

This study represents the largest analysis of SDEC services to date, providing evaluation at both patient and unit level that has not been previously reported. There are currently no other multicentre studies evaluating the delivery of SDEC, and the data reported here is not available through any routinely collected data.(38) There are approximately 250 AMUs within the UK, although the reported number varies and fluctuates, in part due to frequent changes such as mergers between hospital sites;(39) 80% of services contributed data that has been included in this analysis, however an estimated 57% of units provided detailed information describing unit structure in SAMBA22. There may be systematic differences in those hospitals that did not participate; the higher response rate from English services (Supplementary Table 1) means the results may be less reflective of practice in the other nations, which have different policy approaches to SDEC.(24,40) Our data represents a single day within each year, and variation in performance may be expected across time.

All data regarding availability of services and unit structure was self-reported, and therefore at risk of bias. We assume that if the clinical team are unaware of pathways and standard procedures available, then they were not being utilised. Not all suggested standards were evaluated; additional metrics including clinician assessment of patients within an hour of arrival to SDEC, utilisation of validated risk stratification tools for specific conditions, and regular review of SDEC performance using pre-defined metrics should be evaluated in future audit.

Our analysis did not show any significant difference in the provision of hospital services when stratifying by hospital size. For the purposes of our analysis presented here, hospitals with less than 400 beds were grouped as ‘smaller’,(41) due to the low number of small hospitals in this sample. There may be differences in the delivery of care at hospitals that are small or rural,(42) and these hospitals have different demands, access to specialist services and pathways, and logistic differences such as patient travel time, that may affect how SDEC is delivered. Further in-depth evaluation of how SDEC services currently function in these specific settings may provide helpful insights into effective operation when influenced by these factors but requires an alternative methodology to that reported here.

Further focussed research is needed to ensure effective delivery of medical SDEC, and equity of access across hospitals and patient cohorts. This will necessitate robust studies of multiple aspects of service organisation, alongside prospective evaluation of outcomes for patients assessed in SDEC, building an evidence base for medical SDEC which can inform more comprehensive guidance & policy from key groups, including SAM and the NHS.

## Conclusion

Medical same day emergency care services continue to be a key component of assessment pathways within acute medical services, with a third of unplanned medical attendances managed through these pathways. There is considerable variation in provision of these services nationally, in both operational factors and clinical delivery. Further evaluation is needed to understand how SDEC services can be more effectively delivered across different patient populations and hospital settings, and to ensure patients can receive care within the most appropriate setting.

## Author contributions

CA and MP designed and conducted data analysis and drafted the initial manuscript. CA, TK, TC, MH, CS, DL and RV contributed to design of initial data collection. All authors contributed to and approved the final manuscript.

## Data Availability

Data are available on reasonable request. Data from this study are available from PIONEER, the Health Data Hub in Acute care, in accordance with Hub processes. See www.pioneerdatahub.co.uk and contact for more details.

## Acknowledgements

The authors would like to acknowledge those that have assisted with the running of SAMBA, and those at the sites that participated in SAMBA.

## Patient and public involvement

There was no specific patient or public involvement in this study.

## Data sharing

Data are available on reasonable request. Data from this study are available from PIONEER, the Health Data Hub in Acute care, in accordance with Hub processes.

See www.pioneerdatahub.co.uk and contact for more details.

