## Supplementary tables and figures for "Provision of medical Same Day Emergency Care services within the UK: analysis from the Society for Acute Medicine Benchmarking Audit"

**Supplementary Table 1: Participation in SAMBA by UK nation**

| | Overall | England | Scotland | Wales | Northern Ireland |
|---|---|---|---|---|---|
| Estimated eligible units | 247 | 190 | 30 | 15 | 12 |
| Participation in any one or more data collection year | 197 (79.8%) | 168 (88.4%) | 12 (40%) | 10 (66.6%) | 7 (58.3%) |
| SAMBA22 unit structure questions | 140 (56.6%) | 122 (64.2%) | 8 (26.7%) | 6 (40%) | 4 (33.3%) |

**Supplementary Table 2:** Compliance with standards for SDEC, with comparison by hospital size. SOP: Standard Operating Procedure. *Chi square test comparing proportion with vs without SOP.

| | **Overall** (N = 139) | **Smaller Hospitals** (N = 40) | **Medium Hospitals** (N = 47) | **Larger Hospitals** (N = 52) | **P value** |
|---|---|---|---|---|---|
| | *n (%)* | *n (%)* | *n (%)* | *n (%)* | |
| **Consultant physically available** | 89 (64) | 30 (75) | 29 (62) | 30 (58) | 0.21 |
| **Nominated clinician for overall leadership** | 118 (85) | 32 (80) | 40 (85) | 46 (88) | 0.53 |
| **Contact non-attenders** | 110 (80) | 28 (72) | 37 (79) | 45 (87) | 0.22 |
| **Collect patient feedback** | 114 (82) | 29 (73) | 43 (91) | 42 (81) | 0.07 |
| **Private area available** | 126 (91) | 36 (90) | 44 (94) | 46 (88) | 0.67 |
| **Presence of a SOP** | | | | | |
| ***Yes*** | 113 (81) | 31 (78) | 38 (81) | 44 (85) | 0.68* |
| ***No*** | 17 (12) | 8 (20) | 5 (11) | 4 (8) | |
| ***Unsure*** | 9 (6) | 1 (2) | 4 (9) | 4 (8) | |

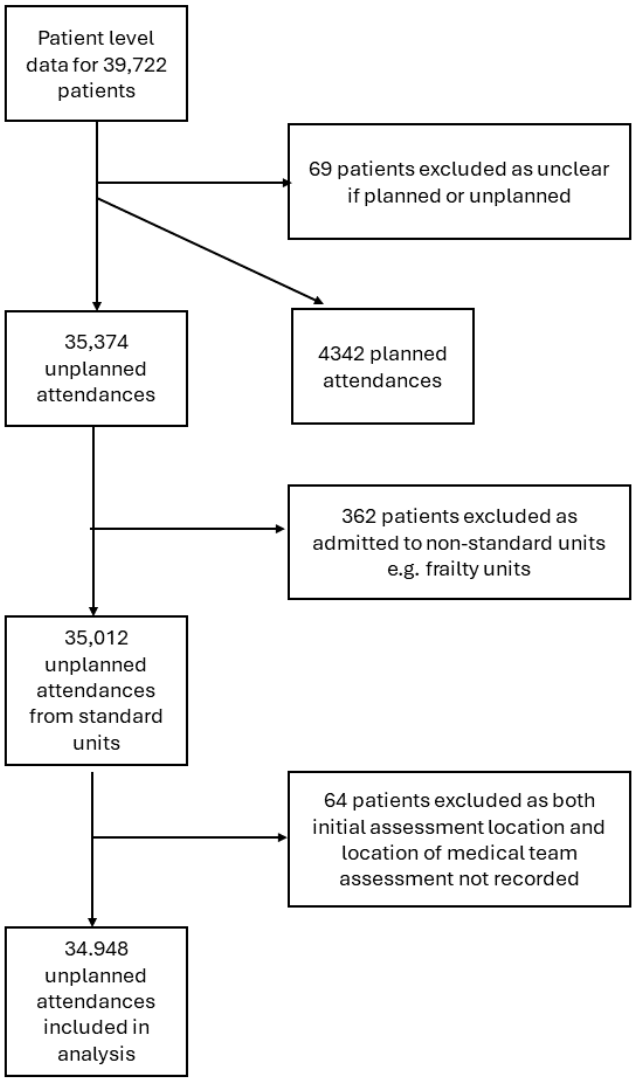


**Supplementary Figure 1:** Flow diagram showing process of exclusion for analysis of patient-level data.

**Supplementary Table 3:** Location of first assessment by the medical team, for unplanned medical attendances by year. SDEC: Same Day Emergency Care; ED: Emergency Department; AMU: Acute Medical Unit.

| | **SDEC** | | **ED** | | **AMU** | | **Other** | | **Total** |
|---|---|---|---|---|---|---|---|---|---|
| | % | *N* | % | *N* | % | *N* | % | *N* | *N* |
| **2019** | 22.1% | *1426* | 37.7% | *2431* | 36.6% | *2362* | 3.6% | *233* | *6452* |
| **2020*** | 21.6% | *1116* | 44.0% | *2274* | 31.1% | *1606* | 3.3% | *169* | *5165* |
| **2021** | 22.2% | *1797* | 40.8% | *3301* | 33.2% | *2684* | 3.7% | *301* | *8083* |
| **2022** | 24.1% | *1724* | 52.2% | *3730* | 21.7% | *1551* | 1.9% | *138* | *7143* |
| **2023** | 29.8% | *2403* | 48.5% | *3903* | 19.2% | *1542* | 2.6% | *206* | *8054* |

| | | SAMBA19 | | Winter SAMBA20 | | SAMBA21 | | SAMBA22 | | SAMBA23 | | |
|---|---|---|---|---|---|---|---|---|---|---|---|---|
| **Patient population** | | *N* | *%* | *N* | *%* | *N* | *%* | *N* | *%* | | | *P value* |
| Age (years) | *16-29* | 157 | 11.1% | 124 | 11.1% | 247 | 13.8% | 192 | 11.1% | 295 | 12.3% | 0.002 |
| | *30-39* | 155 | 10.9% | 139 | 12.5% | 244 | 13.6% | 204 | 11.8% | 321 | 13.4% | |
| | *40-49* | 186 | 12.0% | 163 | 14.6% | 273 | 15.2% | 229 | 13.3% | 299 | 12.4% | |
| | *50-59* | 257 | 18.0% | 178 | 16.0% | 275 | 15.3% | 296 | 17.2% | 369 | 15.4% | |
| | *60-69* | 224 | 15.7% | 187 | 16.8% | 256 | 14.3% | 282 | 16.4% | 269 | 15.4% | |
| | *70-79* | 155 | 17.9% | 172 | 15.45 | 287 | 16.0% | 288 | 16.7% | 412 | 17.2% | |
| | *80-89* | 160 | 11.2% | 132 | 11.8% | 178 | 9.9% | 199 | 11.5% | 282 | 11.8% | |
| | *90+* | 32 | 2.2% | 21 | 1.9% | 37 | 2.1% | 34 | 2.0% | 56 | 2.3% | |
| | >70 years | 447 | 31.4% | 325 | 29.1% | 502 | 27.9% | 521 | 30.2% | 750 | 31.2% | 0.137 |
| Gender | *Female* | 826 | 57.9% | 671 | 60.1% | 1026 | 57.1% | 979 | 56.8% | 1358 | 56.5% | 0.267 |
| Care home resident *(missing=9)* | | 17 | 1.2% | 11 | 1.0% | 18 | 1.0% | 12 | 0.7% | 23 | 1.0% | 0.714 |
| Discharged from hospital in preceding 30 days *(missing:9)* | | 159 | 11.2% | 141 | 12.7% | 237 | 13.2% | 229 | 13.3% | 366 | 15.2% | 0.008 |
| Arrival to hospital* | | | | | | | | | | | | 0.49 |
| | *00:00-08:00* | 39 | 2.7% | 36 | 3.2% | 62 | 3.5% | 100 | 5.8% | 140 | 5.8% | |
| | *08:00-20:00* | 1366 | 95.8% | 1058 | 94.8% | 1715 | 95.4% | 1586 | 92.0% | 2222 | 92.5% | |
| | *20:00-23:59* | 21 | 1.5% | 22 | 2.0% | 20 | 1.1% | 38 | 2.2% | 41 | 2.2% | |
| NEWS* | | | | | | | | | | | | <0.005 |
| 0 | | 858 | 60.9% | 594 | 53.6% | 1052 | 58.5% | 990 | 57.6% | 1133 | 53.6% | |
| 1 | | 327 | 23.2% | 287 | 25.9% | 478 | 26.6% | 435 | 25.3% | 584 | 27.6% | |
| 2 | | 113 | 8.0% | 120 | 10.8% | 157 | 8.7% | 167 | 9.7% | 230 | 10.9% | |
| 3 | | 58 | 4.1% | 64 | 5.8% | 60 | 3.3% | 76 | 4.4% | 100 | 4.7% | |
| 4 | | 25 | 1.8% | 22 | 2.0% | 28 | 1.6% | 28 | 1.6% | 44 | 2.1% | |
| 5 | | 9 | 0.6% | 14 | 1.3% | 13 | 0.7% | 15 | 0.9% | 10 | 0.5% | |
| 6 | | 12 | 0.9% | 4 | 0.4% | 7 | 0.4% | 5 | 0.3% | 8 | 0.4% | |
| 7+ | | 6 | 0.4% | 4 | 0.4% | 2 | 0.1% | 4 | 0.2% | 4 | 0.2% | |
| *Missing* | | 18 | | 7 | | 0 | | 4 | | 290 | | |
| NEWS ≥3 | | 110 | 7.8% | 108 | 9.7% | 110 | 6.1% | 128 | 7.4% | 126 | 7.9% | 0.01^ |
| CFS (in over 70s) (n=1720) | | (n/a) | (n/a) | (325) | | (409) | | (369) | | (617) | | 0.001 |
| 1 | | | | 45 | 13.9% | 39 | 9.5% | 44 | 11.9% | 38 | 6.2% | |
| 2 | | | | 56 | 17.2% | 68 | 16.6% | 71 | 19.2% | 88 | 14.3% | |
| 3 | | | | 84 | 25.9% | 105 | 25.7% | 119 | 32.3% | 170 | 27.6% | |
| 4 | | | | 55 | 16.9% | 92 | 22.5% | 62 | 16.8% | 140 | 22.7% | |
| 5 | | | | 34 | 10.5% | 42 | 10.3% | 43 | 11.7% | 87 | 14.1% | |
| 6 | | | | 24 | 7.4% | 44 | 10.8% | 19 | 5.2% | 66 | 10.7% | |
| 7 | | | | 21 | 6.5% | 13 | 3.2% | 9 | 2.4% | 23 | 3.7% | |
| 8 | | | | 5 | 1.5% | 2 | 0.5% | 2 | 0.5% | 4 | 0.7% | |
| 9 | | | | 1 | 0.3% | 4 | 1.0% | | | 1 | 0.2% | |
| CFS ≥5 | | | | 85 | 26.2% | 105 | 25.7% | 73 | 19.8% | 181 | 29.3% | 0.012 |

**Supplementary Table 4**: Patient characteristics for unplanned attendances receiving their medical team assessment within SDEC services, compared by year. Data for 8466 unplanned attendances.

| | **SDEC** | | **ED** | | **AMU** | | **Other** | |
|---|---|---|---|---|---|---|---|---|
| | % | *n* | % | *n* | % | *n* | % | *n* |
| Discharged without overnight admission | 82.4% | *6908* | 12.8% | *1980* | 15.2% | *1469* | 21.3% | *223* |
| Discharged on day 1-7 | 11.3% | *944* | 49.5% | *7676* | 52.2% | *5053* | 43.6% | *456* |
| In-hospital at day 8 - continuous stay | 4.4% | *366* | 29.5% | *4578* | 26.5% | *2572* | 27.2% | *284* |
| In-hospital at day 8 - readmitted after discharge | 0.6% | *51* | 0.9% | *133* | 0.8% | *77* | 1.1% | *11* |
| Transferred to other healthcare facility | 0.3% | *27* | 1.9% | *298* | 1.7% | *161* | 1.5% | *16* |
| Died in hospital | 0.1% | *6* | 3.6% | *558* | 2.4% | *229* | 1.8% | *19* |
| Self-discharged | 0.9% | *78* | 1.8% | *282* | 1.3% | *129* | 3.5% | *37* |

**Supplementary Table 5:** Outcomes after 7 days for unplanned attendances, by location of assessment by the medical team. SDEC: Same Day Emergency Care; ED: Emergency Department, AMU: acute medical unit.

| | OR | P value | 95% CI |
|---|---|---|---|
| Age | | | |
| 16-29 | *Reference* | | |
| 30-39 | 0.89 | 0.065 | 0.79-1.01 |
| 40-49 | 0.81 | <0.005 | 0.72-0.90 |
| 50-59 | 0.62 | <0.005 | 0.55-0.69 |
| 60-69 | 0.47 | <0.005 | 0.42-0.52 |
| 70-79 | 0.33 | <0.005 | 0.30-0.37 |
| 80-89 | 0.22 | <0.005 | 0.20-0.24 |
| 90+ | 0.17 | <0.005 | 0.15-0.20 |
| Gender | | | |
| Female | *Reference* | | |
| Male | 0.87 | <0.005 | 0.82-0.92 |
| Other | 0.78 | 0.745 | 0.18-3.45 |
| Care home residence | | | |
| No | *Reference* | | |
| Yes | 0.56 | <0.005 | 0.48-0.66 |
| Recent discharge | | | |
| No | *Reference* | | |
| Yes | 0.68 | <0.005 | 0.63-0.73 |
| NEWS score | 0.69 | <0.005 | 0.68-0.70 |
| Time of arrival | | | |
| 00:00-07:59 | 1.03 | 0.502 | 0.95-1.11 |
| 08:00-19:59 | *Reference* | | |
| 20:00-23:59 | 0.25 | <0.005 | 0.22-0.28 |
| Source of referral | | | |
| ED | *Reference* | | |
| 111 | 1.07 | 0.688 | 0.76-1.51 |
| GP | 2.88 | <0.005 | 2.71-3.06 |
| Other hospital | 0.97 | 0.793 | 0.75-1.24 |
| Own hospital (OPD) | 1.96 | <0.005 | 1.67-2.29 |
| Own hospital (other) | 2.26 | <0.005 | 1.91-2.68 |
| Paramedic | 1.10 | 0.238 | 0.94-1.30 |
| Year | | | |
| 2019 | *Reference* | | |
| 2020 | 1.08 | 0.088 | 0.99-1.19 |
| 2021 | 1.24 | <0.005 | 1.14-1.35 |
| 2022 | 1.12 | 0.007 | 1.03-1.22 |
| 2023 | 1.31 | <0.005 | 1.21-1.43 |

**Supplementary Table 6:** Logistic regression model for likelihood of same day discharge. Pseudo $R2=0.1949$. OR: odds ratio; NEWS: National Early Warning Score; ED: Emergency Department; GP: general practice; OPD: outpatient department.

| Row Labels | SAMBA19 | Winter SAMBA20 | SAMBA21 | SAMBA22 | SAMBA23 |
|---|---|---|---|---|---|
| Discharged without overnight admission | 80.1% | 81.8% | 85.3%* | 81.4% | 82.8% |
| Discharged on day 1-7 | 12.8% | 13.3% | 8.7% | 11.5% | 11.2% |
| In-hospital at day 8 - continuous stay | 4.5% | 3.5% | 4.1% | 5.2% | 4.4% |
| In-hospital at day 8 - readmitted after discharge | 1.1% | 0.7% | 0.6% | 0.5% | 0.4% |
| Transferred to other healthcare facility | 0.3% | 0.2% | 0.3% | 0.5% | 0.3% |
| Died in hospital | 0.07% | 0.09% | 0.11% | 0.06% | 0.04% |
| Self-discharged | 1.2% | 0.5% | 1.0% | 0.9% | 0.9% |

**Supplementary Table 7:** Outcomes at day 8 for unplanned attendances receiving medical assessment in SDEC, by year.
